# Trends in HIV self-testing uptake in Africa: a modeling study of population-based surveys and HIV testing program data

**DOI:** 10.1101/2025.09.22.25336306

**Authors:** Aishi Aratrika, Carla M. Doyle, Cheryl Case Johnson, Olanrewaju Edun, Bogol Mbope Mweng, Achille Adoko, Mphotleng Tlhomola, Cédric P. Yansouni, Augustine Talumba Choko, Jeffrey W. Imai-Eaton, Mathieu Maheu-Giroux

**Affiliations:** Department of Epidemiology and Biostatistics, McGill University, Montréal, Québec, Canada; HIV, Hepatitis and STI Department, World Health Organization, Geneva, Switzerland; MRC Centre for Global Infectious Disease Analysis, School of Public Health, Imperial College London, London, United Kingdom; Ministry of Health, Kinshasa, Democratic Republic of the Congo; The Joint United Nations Program on HIV/AIDS (UNAIDS) Country Office, Benin; Ministry of Health, Maseru, Lesotho; JD MacLean Centre for Tropical and Geographic Medicine, and Divisions of Infectious Diseases and Medical Microbiology, McGill University Health Centre, Montréal, Québec, Canada; Malawi Liverpool Wellcome Trust Clinical Research Programme, Blantyre, Malawi; Center for Communicable Disease Dynamics, Department of Epidemiology, Harvard T.H. Chan School of Public Health, Harvard University, Boston, United States

**Keywords:** HIV self-testing, HIV testing services, HIV, AIDS, mathematical modeling, Africa

## Abstract

**Background:** HIV self-testing (HIVST) can increase access to and uptake of HIV testing among people underserved by other HIV testing approaches. Several countries in Africa, the region most affected by HIV, have scaled-up HIVST. We aimed to estimate 1) country-level and regional trends in HIVST uptake among adults by sex and age and 2) the proportion of distributed HIVST kits that are used and re-testing rates with HIVST.

**Methods:** Across African countries, we analyzed 1) data from national population-based surveys that included questions on previous HIVST use and 2) the number of HIVST kits distributed from nationally reported program data (2012-2024). We developed a hierarchical Bayesian compartmental model to estimate HIVST rates by triangulating surveys and program data. Random effects were used to pool information across countries.

**Results:** Data were available from 40 surveys in 27 countries and from 99 country-years of HIVST program data. The proportion of adults aged ≥15 years in Africa who have ever used an HIVST (HIVST uptake) steadily increased, from <1% in 2012 to almost 7% (6.8%; 95% credible interval [95%CrI]: 5.8%–8.2%) in 2024. HIVST uptake was higher in eastern and southern Africa (10.2% in 2024, 95%CrI: 8.5%-12.7%) compared to western and central Africa (2% in 2024; 95%CrI: 1.7%-2.5%) The proportion of people who ever self-tested varied substantially across countries, reaching a maximum in 2024 of 45.4% (95%CrI: 41.8%-51.5%) in Lesotho. Men (7.2% in 2024, 95%CrI: 6.1%–8.8%) were more likely than women to have ever used an HIVST (6.4% in 2024; 95%CrI: 5.4%-7.8%). Compared to younger individuals (15-24 years), those aged 25–34 years had higher rates of self-testing (men: rate ratio[RR]=1.8, 95%CrI: 1.5-2.3; women: RR=1.4, 95%CrI: 1.1-1.6). Individuals who previously self-tested had a 10% higher probability (RR=1.1, 95%CrI: 0.8-1.5) to self-test again. We estimated that 70% (95%CrI: 60%-80%) of all HIVST distributed were used.

**Interpretation:** HIVST uptake has increased in Africa, with wide between-country variations. Compared to other testing modalities, HIVST is more likely to engage older-to-midlife adults and men who have historically been less likely to be aware of their HIV status. Our results can help understand patterns of use and support countries in optimizing their testing services.

**Funding:** Canadian Institutes of Health Research, Canada Research Chairs, Children’s Investment Fund Foundation.

## Introduction

HIV testing services (HTS) are essential for reaching people living with HIV (PLHIV) who do not know their status and are a critical entry point to both HIV prevention and treatment services. Despite the substantial success scaling-up HTS over the last three decades, gaps in testing coverage remain [1]. HIV self-testing (HIVST), whereby an individual performs a blood-based or oral rapid test and interprets the result on their own in private settings, has been shown to be an important tool that can help address remaining gaps by increasing access to and uptake of HIV testing [2,3].

Africa remains disproportionately affected by HIV, home to 26 million PLHIV and experiencing an estimated 640,000 new HIV acquisitions in 2023 [4]. To end the HIV epidemic, the *Global AIDS Strategy* has set targets for 95% of PLHIV to be aware of their status, 95% of diagnosed PLHIV on treatment, and 95% of treated PLHIV to be virally suppressed by 2030 [5]. Countries in eastern and southern Africa are closer (around 93% PLHIV being aware of their status in 2023) to achieving this 95% diagnosis coverage target, compared to 81% of PLHIV in western and central Africa [6,7].

Progress toward the first 95 target is hampered by persistent and multifaceted barriers to HIV testing, often disproportionately impacting adult men and key populations. Such barriers include differential engagement with health services that provide less entry points for testing, internalized and anticipated stigma, discrimination, criminalization, punitive and restrictive laws, privacy concerns, gender norms, poverty and geographical inaccessibility [8–15]. While traditional provider-delivered HTS approaches in antenatal care, outpatient settings and focused community outreach have been impactful, closing the remaining gaps will require a more diverse and expanded package of HTS options, particularly for populations with undiagnosed PLHIV who are not accessing services [3,16].

HIVST is a safe, acceptable and discreet HIV testing option to overcome some of the persistent barriers to service delivery. The *World Health Organization* (WHO) first recommended HIVST in 2016 [3,8,17]. Two large-scale multi-country initiatives, the *STAR (HIV Self-Test Africa)* Initiative launched in 2015 and the *ATLAS (Auto Test VIH, Libre d’Accéder à la connaissance de son Statut)* launched in 2018, contributed to expanding self-testing and HIV diagnosis in distinct epidemiological contexts [18,19]. Mathematical modeling and economic evaluations based on these programs demonstrated that HIVST improve treatment coverage, reduce HIV incidence, and are consistently cost-effective [20,21]. Routinely monitoring HIVST uptake and its subsequent effects is challenging, however. Due to its private and flexible nature, program data typically capture only the number of HIVST kits distributed, without information on actual use, user characteristics, confirmed test results, or onward linkage to further testing, treatment or prevention services. Disclosure of reactive self-test results may not be consistently reported or recorded at the individual or program level. Population-based surveys collecting self-reported information on awareness and use of HIVST can be useful to track uptake, but are conducted at five-year intervals in the best of cases. Consequently, estimates of HIVST uptake remain limited to country-specific, small-scale studies on HIVST trends [22–26].

To improve our understanding of HIVST uptake, we synthesized self-reported HIVST uptake data from population-based surveys and annual self-test kit distribution data across African countries. We developed a hierarchical mathematical model to triangulate these data sources and generate country and regional estimates of HIVST uptake by sex and adult age groups. We also estimated HIVST re-testing rates and the proportion of HIVST kits that were likely used from the model. Lastly, to assess whether HIVST uptake varies by HIV status, we conducted a separate random-effects meta-analysis using only survey data.

## Methods

### Data sources

We collated data from two sources: *(1)* nationally representative population-based surveys that included questions on participants’ self-reported previous use of HIVST and *(2)* country-level HIV testing program data on the annual number of HIVST kits distributed. To identify eligible surveys, we reviewed all publicly available, cross-sectional, population-based surveys from countries across eastern, southern, central and western Africa conducted between January 2012 to December 2024. The earliest date corresponds to the first survey to include self-reported HIVST uptake information, which was the 2012 *Kenya AIDS Indicator Survey* (KAIS) [27]. We systematically searched data catalogs (i.e*., the Global Health Data Exchange, the WHO Multi-Country Studies Data Archive*) and conducted Google engine and literature searches. We identified the surveys with HIVST usage-related items, often measured through the question ‘*Have you ever tested yourself for HIV using a self-test kit?’* (see Table S1.1 for list of all survey questions). The following survey series were identified: *Demographic and Health Surveys* (DHS), *AIDS Indicator Cluster Surveys* (AIS), *Multiple Indicator Cluster Surveys* (MICS), *Population-based HIV Impact Assessments* (PHIA), *Botswana AIDS Impact Survey* (BAIS) and *Kenya AIDS Indicator Survey* (KAIS). For each survey, we calculated the sex-and age-stratified proportions of respondents aged ≥15 years who reported ever using an HIVST, applying the survey weights accounting for the multi-stage cluster sampling designs.

We collected information on the annual number of HIVST kits distributed between January 2016 and December 2024 from multiple sources: Spectrum model files submitted by national HIV programs to UNAIDS for the 2024 national HIV estimates round, the *WHO/UNAIDS Global AIDS Monitoring* (GAM) system (2018-2023), the 2024 *PANAROMA* dataset of the *President’s Emergency Plan for AIDS Relief* (PEPFAR), and national HIVST procurement records reported to WHO (2016-2024). For both Spectrum and GAM, HIV testing data are routinely reported jointly to the WHO and the UNAIDS by countries through their national HIV testing programs. Where data for the same country-year were available from multiple sources, we prioritized data in the following order: 1) Spectrum estimates files, 2) GAM, or 3) PEPFAR as Spectrum allows countries to update or revise previously submitted data in subsequent years, whereas data submitted to GAM are fixed once reported and cannot be modified retrospectively.

Our analysis considered countries with at least one available survey and one year of HIVST distribution data, as these were the minimum data required to calibrate the model.

### Mathematical modeling of HIVST uptake

We developed a novel deterministic compartmental model to estimate HIVST uptake over 2012-2024 among open populations of individuals aged +15 years based on a system of ordinary differential equations (Figure 1). The model partitioned the population into individuals who have never used an HIVST (*N*) and individuals who have ever used an HIVST (*H*). We further stratified the model by sex and age groups (15–24, 25–34, 35–49 and ≥50 years). Entry into the model occurs in the never HIVST compartment for individuals reaching age 15 years and individuals exit the model through death. Demographic inputs were sourced from the United Nations 2024 *World Population Prospects* (WPP) Revision [28].

**Figure 1:**
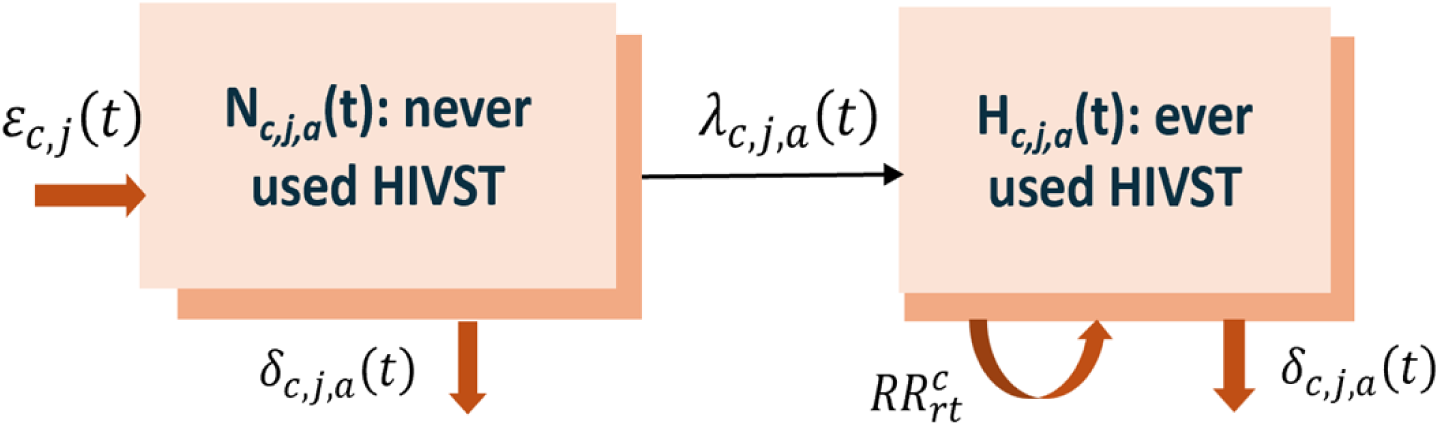
*Diagram of the compartmental flows of the deterministic model of HIV self-test (HIVST) users and non-users.* **Indices: c = country, j = sex, a= age group (15-24, 25-34, 35-49, ≥50), t = year (2012-2024).** 𝜀_𝑐,𝑗_(𝑡): yearly entry rate of 15-24 year-olds; 𝛿_𝑐,𝑗,𝑎_(𝑡): annual death rate; 𝜆_𝑐,𝑗,𝑎_(𝑡): annual HIV self-testing rate; 𝑅𝑅_𝑟𝑡_: HIV self-testing retesting rate ratio.

All individuals were assumed to have never used an HIVST at model initiation in 2012. People can use HIVST at a time-varying rate (λ_𝑐,𝑗,𝑎_(𝑡)) which was modeled as a first-order random walk with annual time steps (*t*). These self-testing rates are country-specific (*c*) and vary by sex (*j*) and age groups (*a*). Individuals who previously used HIVST may be more or less likely to use it again, depending on how HIVST distribution is implemented (e.g., HIVST distributed to key populations). To account for that, we introduced a re-testing rate ratio (RR) for people in the ever-self-tested (*H*) compartment.

The model parameters’ priors are presented in Table 1. Our main assumptions, differential equations, and other additional details can be found in the Supplementary Materials (Section S3).

**Table 1:**
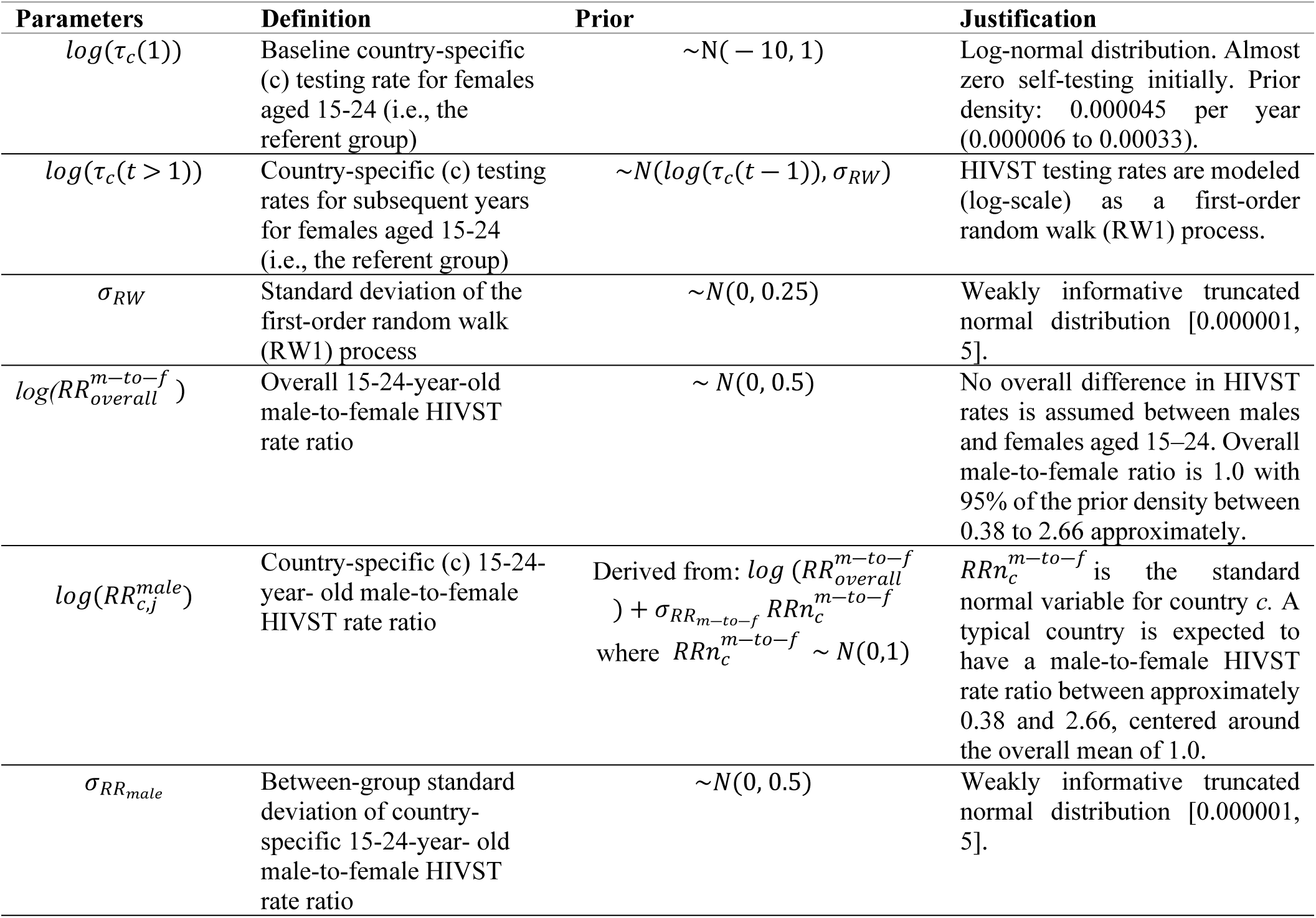
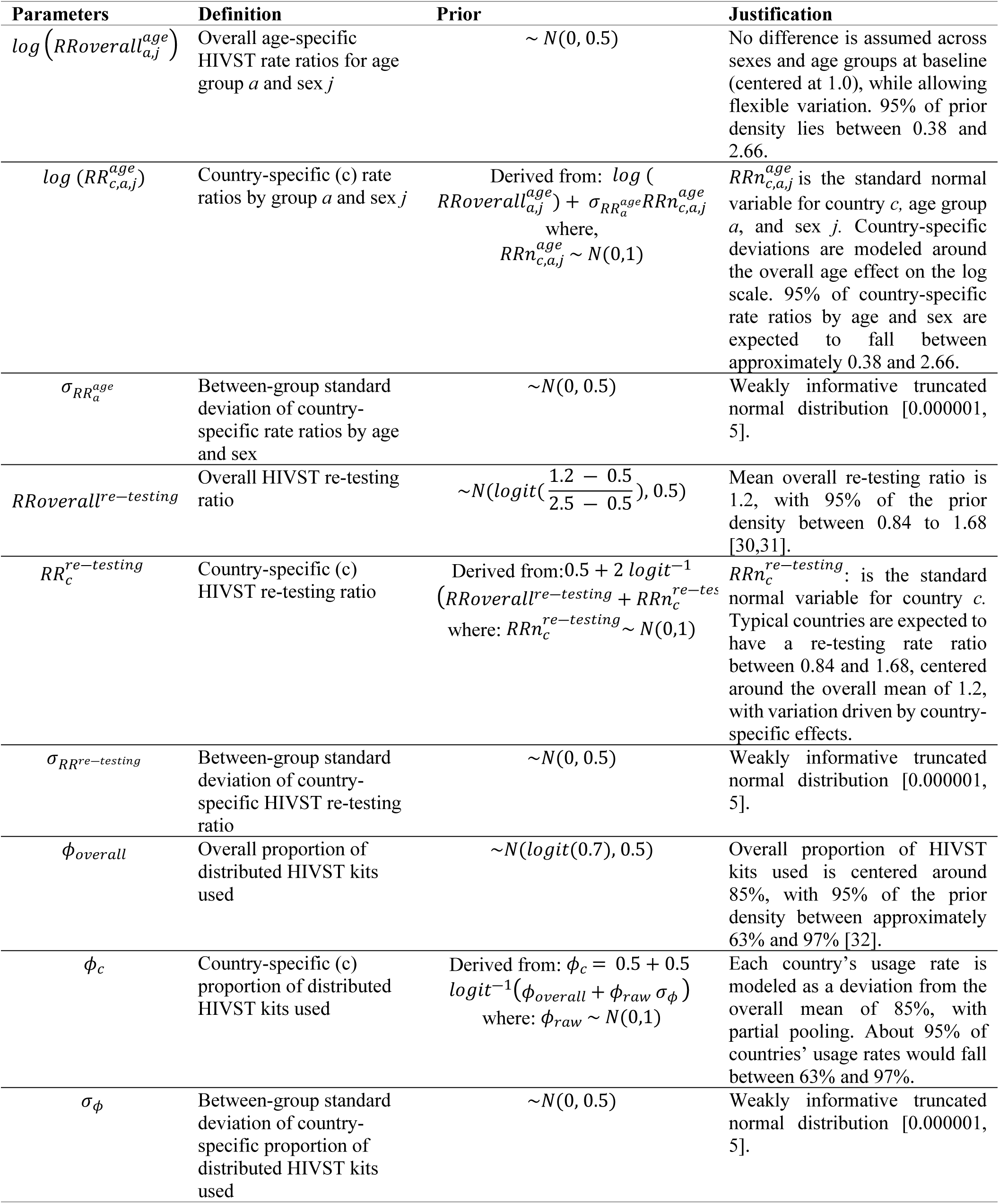
List of main parameters, their definitions, prior distributions, and justifications for the mathematical model of HIV self-testing uptake.

### Model calibration

The model was calibrated using a Bayesian framework with a hierarchical structure on rate ratio parameters, borrowing strength across countries, with weakly informative priors. The calibration outcomes include: *1)* the survey proportion of the population reporting having ever used an HIVST, stratified by sex and age groups (using a binomial likelihood), and *2)* the annual number of HIVST distributed (using a normal distribution likelihood).

We calibrated five parameters per country: the male-to-female RRs for the referent age groups, the sex-specific RRs for each age groups, the proportion of distributed kits used, and the HIVST re-testing RR. The first three parameters are informed by the survey data (as the program data contains no information on sex or age of the users), whereas an overlap between the survey and the program data for a given year informs the proportion of HIVST used and the re-testing ratio. In addition, we incorporated a soft quadratic penalty in years where program data were missing to avoid unrealistically high HIVST testing rates. The penalty threshold was set for the model-predicted annual number of HIVST distributed to twenty times the size of 0.10% of the population for countries with maximum annual testing coverage below 0.10%, and to four times the maximum observed number of HIV tests for all other countries. To obtain posterior distributions for the estimated parameters, we used a Hamiltonian Monte Carlo (HMC) sampling algorithm with four chains and 4,000 iterations each (including 2,000 iterations for warm-up) implemented in the *rstan* package (version 2.32.7) [29]. We assessed convergence through traceplots and the R-hat statistics and summarized posterior distributions by the median and 95% credible intervals (95%CrI).

### Model outputs

Using the calibrated model, we calculated the proportion of the population that ever used an HIVST by country, year, sex, and age group, and the annual total number of HIVST kits used for each country from 2012 to 2024. To calculate the aggregated trends by age groups (14-24, 25-34, 35-49, ≥50 years), sex (men/women) and region (western and central Africa/eastern and southern Africa), the country and year-specific estimates for HIVST uptake were weighted by their relative population size.

### Meta-analysis of HIVST uptake by HIV serostatus

The model above did not stratify the population by HIV serostatus since most surveys did not collect that information. To assess whether HIVST distribution is reaching PLHIV not on treatment, we performed a meta-analysis of self-reported HIVST usage by serostatus. We focused on PLHIV who are not on treatment since those who are do not require being diagnosed again. ART use was based on either a self-report of being on treatment or having biomarkers of antiretroviral drugs. Specifically, we performed logistic regression separately on each survey to estimate the odds ratio of having used an HIVST among people not living with HIV and PLHIV not on treatment. The logistic regression models were adjusted for sex (male, female), geographical region (urban, rural), age group (5-year age groups), household wealth quintile (lowest, second, middle, fourth, highest), and educational attainment (less than primary education, primary education, secondary education, more than secondary education), and accounted for clustering by primary sampling units. Using the *metafor* package, we pooled survey-specific odds ratios with a random-effects meta-analysis based on Restricted Maximum Likelihood (REML).

All the analyses were performed using R software *(version 4.4.3)*. This study complies with the *Guidelines for Accurate and Transparent Health Estimates Reporting* (GATHER) statement (Supplementary Materials, Table S2.1).

### Ethics

All individuals participating in the survey provided verbal informed consent. The protocols for the DHS surveys have been approved by the Internal Review Board of ICF International in Calverton, USA. For all other surveys included in this study (MICS, PHIA, BAIS and KAIS), approval was obtained from relevant country authorities. Ethical approval for secondary data analyses was obtained from the Institutional Review Board of McGill University (A10-E72-17B).

## Results

### Included population-based surveys and countries with HIVST program data

We reviewed 93 surveys in Africa from 2012 to 2024 and identified 46 surveys from 33 countries with self-reported HIVST usage. Of the surveys identified, there were 23 DHS, 14 MICS, 7 PHIA, 1 BAIS, and 1 KAIS surveys. Among the 33 countries with survey data, we excluded six that did not have any HIVST program data: the Central African Republic, Chad, Comoros, Gabon, Mauritania, and Togo. Among countries with program data, these were available for a median of 4 years, totaling 99 country-years of information on 20 million HIVST kits distributed. The final analysis included 40 surveys conducted in 27 countries containing information on 854,501 respondents aged ≥15 years, with 10 countries having more than one survey (Figure 2).

**Figure 2:**
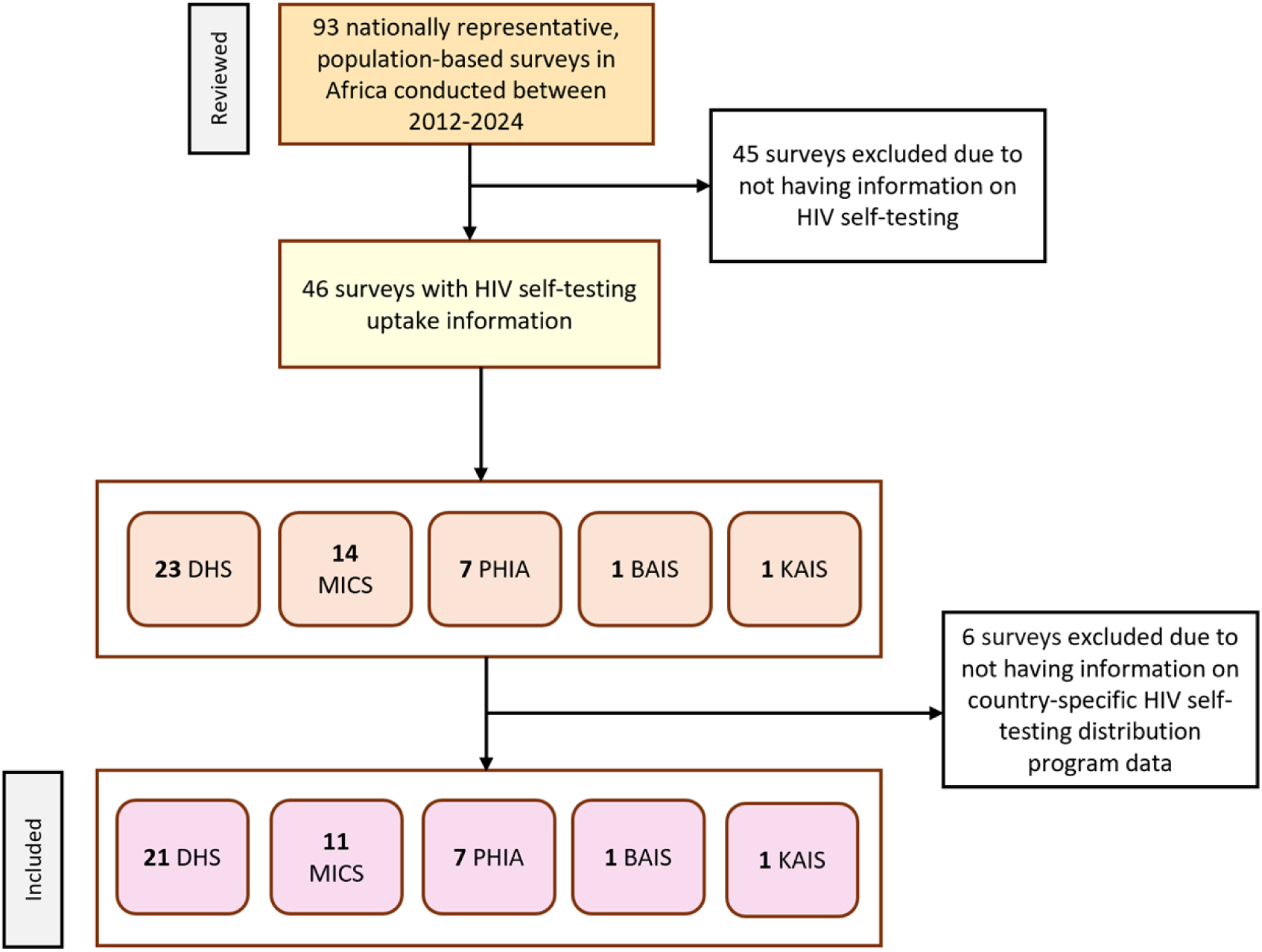
Flowchart of population-based surveys reviewed for inclusion to inform the model estimated proportion of HIV self-testing (*HIVST) uptake in Africa (2012-2024).* DHS: Demographic and Health Survey; MICS: Multiple Indicator Cluster Survey; PHIA: Population-based HIV Impact Survey; BAIS: Botswana AIDS Impact Survey; KAIS: Kenya AIDS Indicator Survey.

### Trends in HIVST uptake by region, sex and age

Our model reliably reproduced HIVST usage estimates from population-based surveys and replicated annual HIVST distribution volumes (Supplementary Materials, Section S4). We estimated that, across the 27 African countries included, the proportion of adults aged ≥15 years who had ever tested for HIV using an HIVST (henceforth defined as HIVST uptake) increased steadily from <1% in 2012 to 6.8% (95%CrI: 5.8%–8.2%) in 2024 (Figure 3A). Although the proportion of HIVST users increased overall, the progress differed by country and region. Before 2016, uptake was similar between regions, but thereafter the countries in western and central Africa had consistently lower uptake (2.0% in 2024; 95%CrI: 1.7%-2.5%) than countries in eastern and southern Africa (10.2% in 2024; 95%CrI: 8.5%-12.7%; Figure 3B).

**Figure 3:**
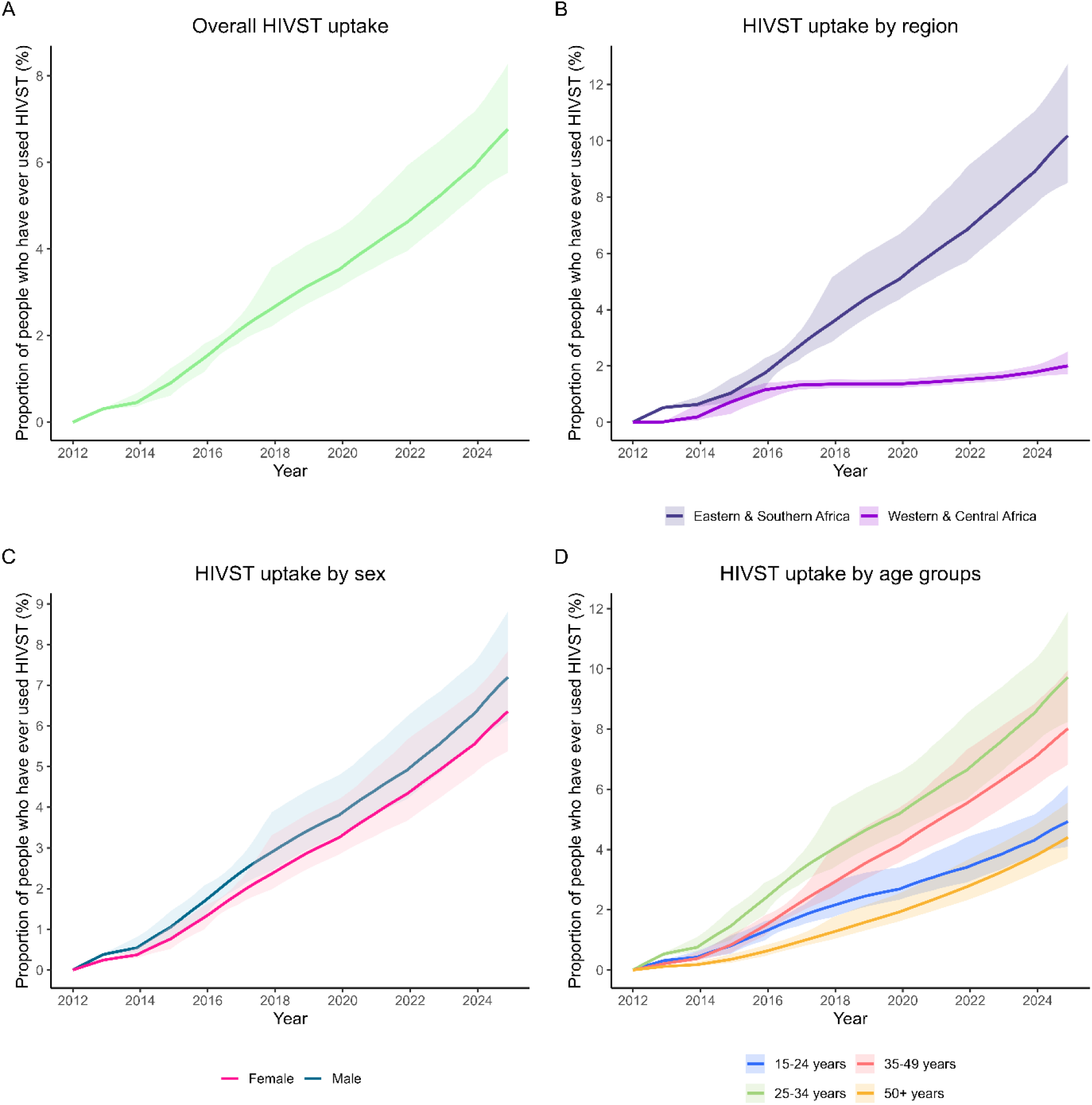
Model estimated trends in HIV self-testing (HIVST) uptake (i.e. proportion who ever used an HIVST) overall (A), by region (B), sex (C) and age groups (D) in Africa (2012-2024). The lines represent the median, and the shaded areas represent the 95%credible intervals.

The uptake of HIVST uptake was slightly higher in men (7.2% in 2024; 95%CrI: 6.1%– 8.8%) than women (6.4% in 2024; 95%CrI: 5.4%-7.8%; Figure 3C). Additionally, in 2024, HIVST uptake was estimated to be highest among 25-34-year-olds (9.7%; 95%CrI: 8.2%–11.9%), followed by 35-49-year-olds (8.0%; 95CrI: 6.8%–10%) and 15-24-year-olds (4.9%; 95%CrI: 4.1%–6%). It was lowest among people aged ≥50 years (4.4%; 95%CrI: 3.7%–5.6%; Figure 3D).

### Country-level estimates of HIVST uptake in 2024

National estimates for the proportion of individuals reporting ever using an HIVST in 2024 were highly heterogeneous (Figure 4). In 2024, eastern and southern African countries had the highest national estimates of HIVST uptake (above 10%): Lesotho (45%; 95%CrI: 42%-51%), Eswatini (30%; 95%CrI: 25%-43%), Malawi (18%; 95%CrI: 15%-30%), Zimbabwe (16%; 95%CrI: 12%-26%), South Africa (12%; 95%CrI: 8%-22%), Namibia (13%; 95%CrI: 10%-24%) and Zambia (11%; 95%CrI: 9%-19%) (Figure 4). In contrast, HIVST uptake as of 2024 was below 1% in the following countries: Madagascar, Benin, Guinea, Senegal, and Burkina Faso, which are countries with low national HIV prevalence (Figure 4).

**Figure 4:**
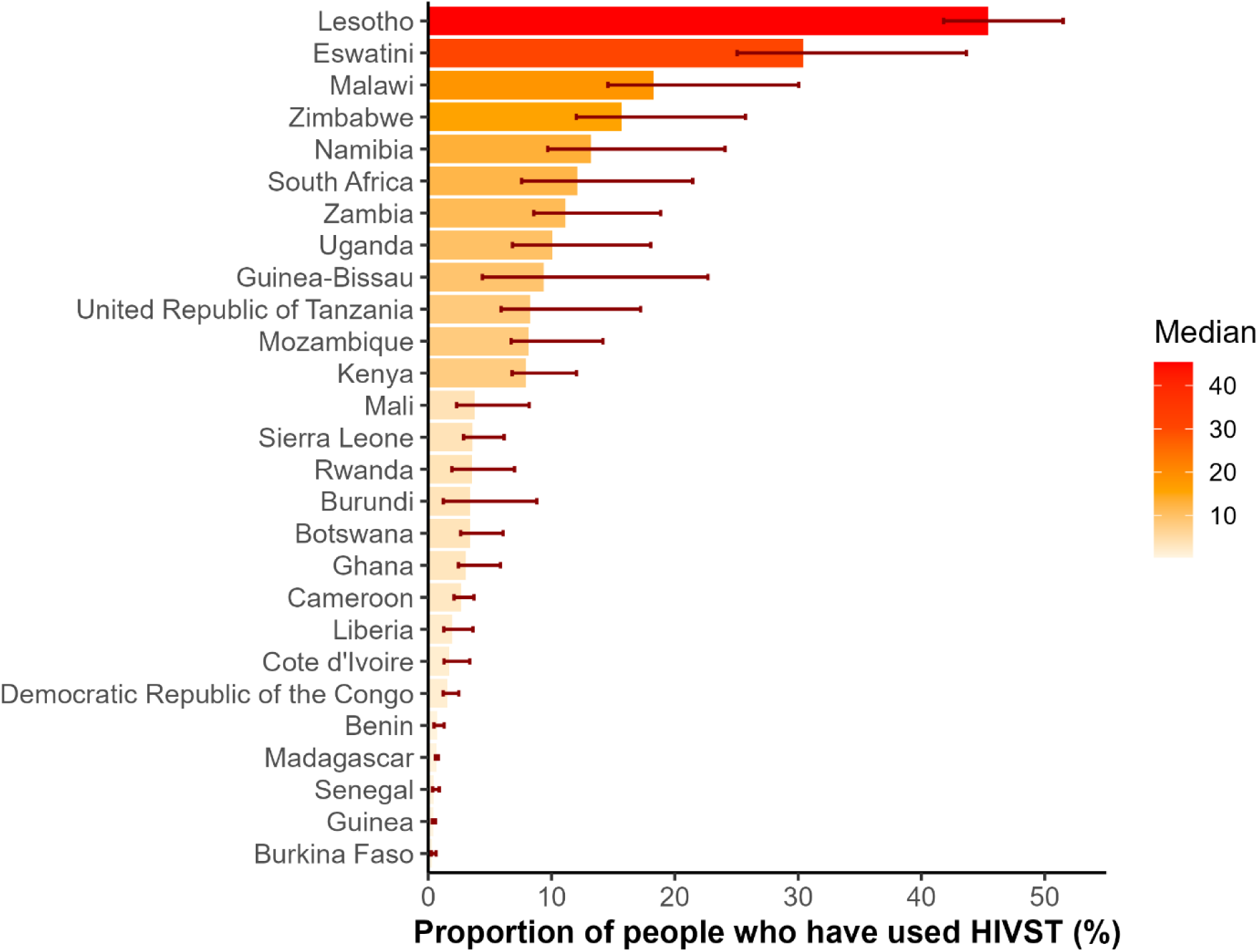
*National estimates of HIV self-testing (HIVST) uptake (i.e., proportion of the population aged 15+ years who ever used an HIVST) in 2024.* The medians represent the posterior median estimates of national HIVST uptake in 2024.

### HIVST rate ratios by age and sex

Among the youngest 15-24-year-olds age group, HIVST rate was lower in men (RR=0.8; 95%CrI: 0.7-0.9) than in women. However, there were substantial variations by country (Figure 5A). For both sexes, the pattern for the age-specific rate ratios was generally consistent across countries (Figure 5B-5C), with 25-34-year-olds testing at higher rates than other age groups. In contrast, people aged ≥35 years had the lowest HIVST rates.

**Figure 5:**
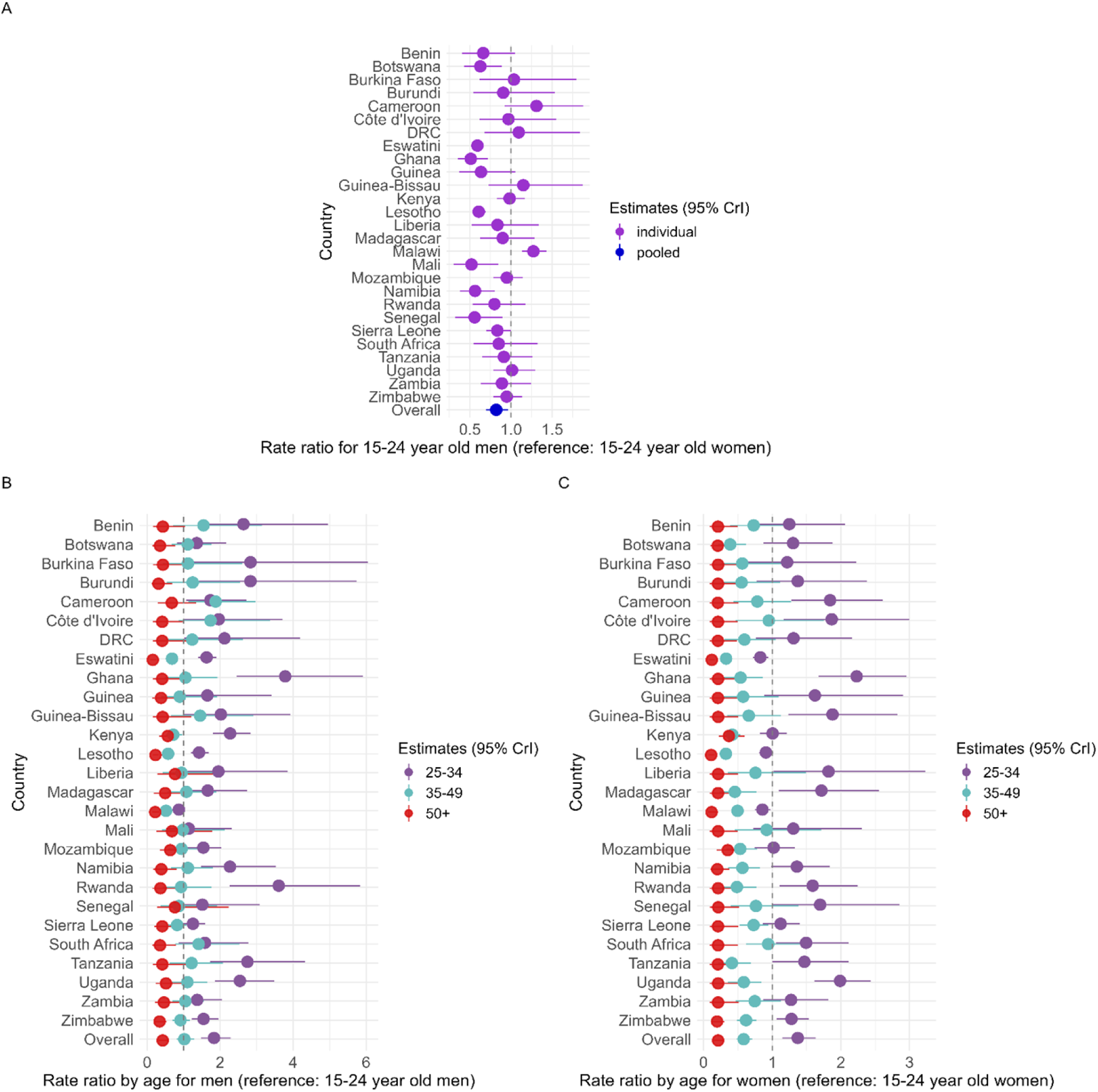
*Overall and country-specific posterior distributions of the following parameters: rate ratios for 15-24-year-old men (A), age-specific rate ratios for men (B), age-specific rate ratios by age group for women (C).* The filled circles represent the median estimates, and the horizontal bars represent the 95% credible intervals (CrI).

The pooled age-specific rate ratios for 25-34-and 35-49-year-olds differed between men and women. By age 25-34 years, men were 1.8 (95%CrI: 1.5-2.3) times more likely to use HIVST than those 15-24-year-olds, while 25-34-years-old women were 1.4 (95%CrI: 1.1-1.6) times more likely than 15-24-year-old women (Figure 5C). In the 35-49-year age group, the difference from the referent group for the overall age-specific rate ratios was less pronounced in men (RR=1.0; 95%CrI: 0.8-1.3; Figure 5B) than in women (RR=0.6; 95%CrI: 0.5-0.7; Figure 5C). Most population-based surveys only collected information on men and women aged 15-49 years, with some surveys including men aged 50+. As such, the rate ratios for that latter group were more precise than those of women.

### HIVST re-testing rate ratios and proportion of distributed HIVST kits used

Compared to first-time HIVST users, we estimated that people who had previously used an HIVST were slightly more likely (RR=1.1; 95%CrI: 0.8-1.5) to re-use one (Figure 6A). However, this parameter was imprecise because an accurate estimate requires a time series of survey and program data that overlap. This information was available from only four countries: Eswatini, Lesotho, Malawi, Mozambique (Figure 6A).

**Figure 6:**
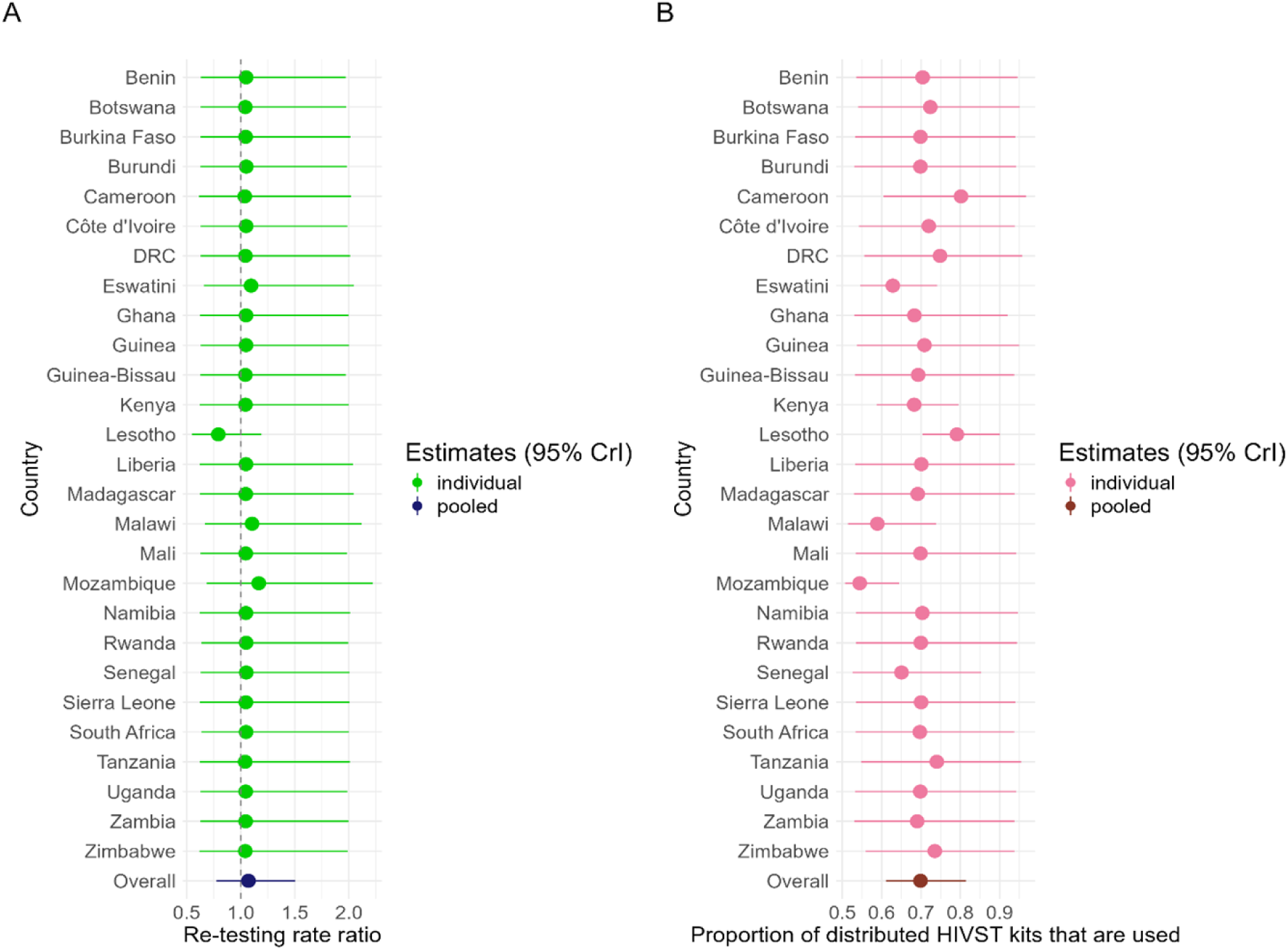
*Overall and country-specific posterior distributions of the following parameters: re-testing rate ratio (A) and proportion of distributed kits used (B).* The filled circles represent the median estimates, and the horizontal bars represent the 95% credible intervals (95%CrI).

The fraction of distributed kits that are used had slightly more information in the data, since it could be estimated when there was a minimum of one year of overlap in years of survey and years of HIV testing program data. Overall, we estimate that 70% (95%CrI: 60%-80%) of distributed HIVST kits are used by end users (Figure 6B). Credible intervals were wide and overlapping for most countries, indicating limited evidence for systematic variation in HIVST usage proportions across countries (Figure 6B).

### HIVST uptake by HIV serostatus

Seventeen surveys with self-reported HIVST usage collected information on HIV serostatus, eight of which had information on ART status (one BAIS and seven PHIAs). Overall, the pooled estimate indicated that PLHIV not on treatment had 25% (odds ratio [OR]=0.75; 95% confidence interval [CI]: 0.53-1.08) lower odds of reporting having used an HIVST as compared to people not living with HIV, after adjusting for sex, age group, urban/rural residence, household wealth, and education. However, there was evidence of moderately high heterogeneity (I^2^=74%; Figure 7). Of the included countries, Kenya (OR=1.84; 95%CI: 1.16-2.92) and Botswana (OR=1.19; 95%CI: 0.48-2.94) had higher odds of HIVST uptake among PLHIV not on treatment, in contrast to the rest of the surveys included in the meta-analysis.

**Figure 7:**
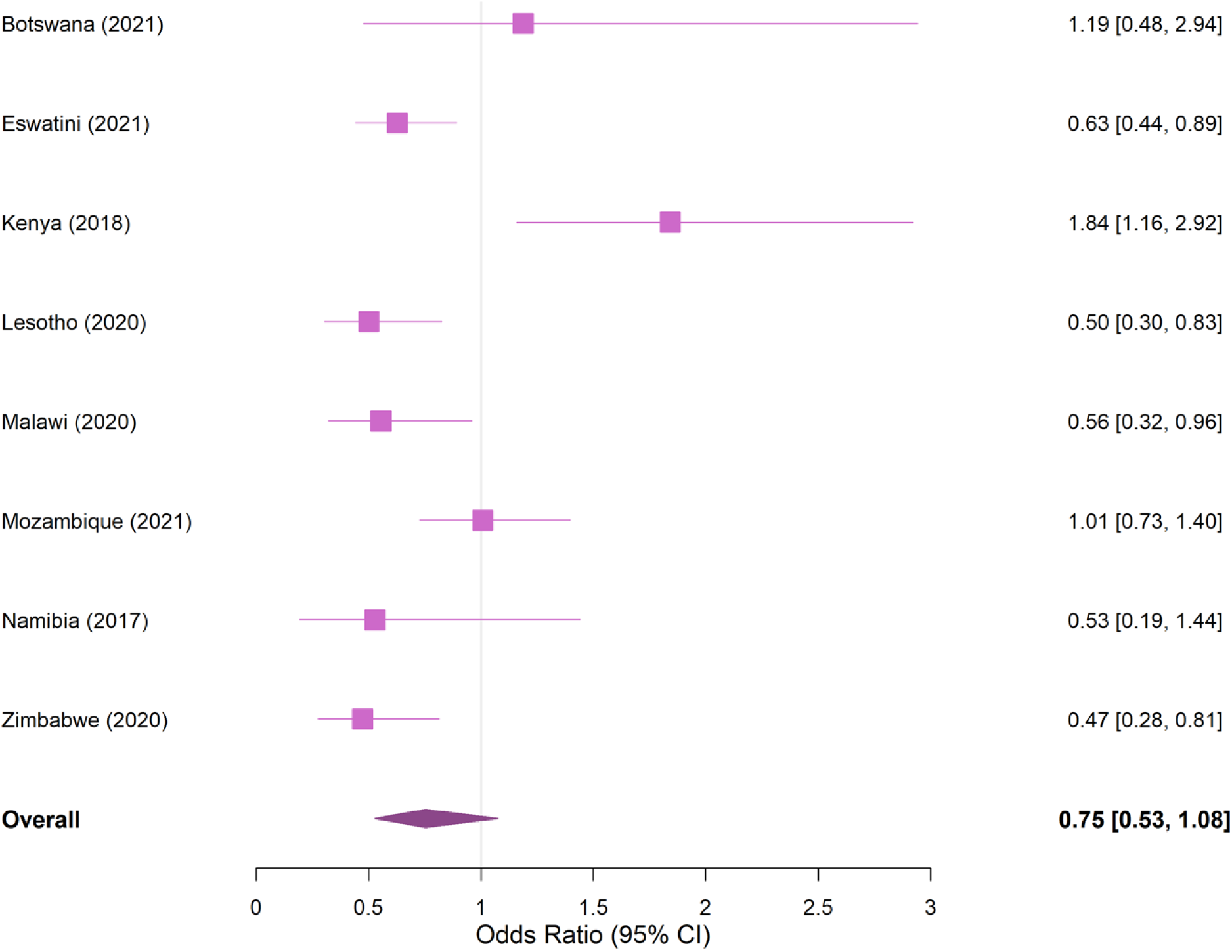
*Forest plot showing the association of HIVST use and HIV serostatus in surveys with available data (BAIS: Botswana, PHIA: Eswatini, Malawi, Mozambique, Kenya, Lesotho, Zimbabwe, Namibia).* Survey-specific odds ratio (OR) estimates and 95% confidence intervals (95%CI) for the logistic regression model predicting HIVST uptake testing among all adults not on ART in 8 countries are indicated by the filled purple squares, and the pooled estimate with its associated 95%CI is shown by the filled purple diamond.

## Discussion

Tracking uptake of HIVST is challenging. Using 40 population-based surveys encompassing more than 850,000 participants and program data from 27 countries, we developed a flexible hierarchical mathematical model of HIVST behaviors. We estimated that as of 2024, 7% of people aged ≥15 years had ever used an HIVST in these countries. Our results suggest that males are more likely to use an HIVST, that individuals aged 25-34 have the highest rates of HIVST use, that about 70% of distributed kits are used, and that individuals who have used an HIVST in the past could be slightly more likely to use it again.

Across Africa, coverage of traditional facility-based HTS has consistently been lower for men compared to women, contributing to consistent gender disparities in HIV testing [33]. A modeling study estimated that men aged 35-49 years were the largest undiagnosed group of PLHIV unaware of their HIV serostatus in 2020 [1,34]. Our results indicate that men are slightly more likely to have used an HIVST (7.2% for men versus 6.4% for women). Such findings are consistent with existing literature [8,35]. Men’s higher uptake of HIVST could be the consequence of existing distribution strategies in Africa that prioritize them such as 1) distribution to partners of pregnant and lactating women; 2) formal and informal workplace distributions; 3) prisons and harm reduction centers; 4) mobile brigades and outreach testing; 5) targeted distribution to key populations, especially men who have sex with men (MSM) and men sex workers [3,36]. This points to the potential role that HIVST can play in closing HIV diagnosis gaps. Lower uptake of HIVST in younger age groups may also be related to policy barriers that limit access due to age of consent laws. While WHO has previously recommended these be reviewed and revised, several countries, particularly in west and central Africa, limit HIV testing and self-testing to those ≥18 years [37].

The scale-up of HIVST was highly variable across countries and regions. The eastern and southern regions, which have higher HIV prevalences, showed greater uptake of HIVST (10%) —five-times that of the western and central regions (2%). Countries where HIVST uptake exceeded 10% in 2024 were all part of the STAR initiative, except Namibia. National HIVST implementation follows two main strategies: 1) distribution prioritized to key populations (e.g., sex workers, MSM), and 2) facility-based distribution through public and private sectors. In practice, however, countries adopt a mix of distribution models [38]. Countries with higher HIV burden and more established ART programs have incorporated HIVST into long-term sustainability plans, using it to reach first-time testers and support routine retesting among vulnerable populations. Irrespective of the strategy adopted, our results indicated that most of the distributed HIVST kits are used, although the pooled estimate has wide uncertainty. Our analysis triangulating survey and program distribution data assumed that HIVST kits were used in the same calendar year they are distributed. This may be inaccurate in situations where kits are intentionally retained for later use or secondary distribution.

Lesotho has the highest estimated HIVST uptake of all included countries. Self-testing in Lesotho has been widely implemented as a screening tool and as a substitute for provider-administered testing [39]. The HIVST delivery model in Lesotho employs both facility-based and community-level distribution models, but it primarily focuses on expanding testing access among the overall population, which likely resulted in our estimated retesting rate ratio of less than one. On the other hand, in Eswatini, facility-based universal screening using HIVST is offered to males aged 20-34 years and females aged 15-24 years, along with peer distribution, community-led distribution, and social network distribution [40]. In this context, the high re-testing rate ratio estimated for Eswatini may be explained by the use of HIVST as a routine tool for specific priority populations and high-risk individuals who could be more likely to engage in frequent self-testing. For Mozambique, the high retesting rate ratio (>1.1) could reflect its key populations-focused HIVST implementation strategy. Within their strategy, there is limited pharmacy-based distribution, but a facility-based distribution of HIVST kits is currently being piloted with a large scale-up planned [41]. In addition, HIVST is used as a complement to conventional (in-facility) tests in Malawi and Zimbabwe, both of which have strong facility-based HIVST implementation and high HIVST uptake. For instance, Malawi incorporated HIVST in over 98% of its facilities [42].

Most western and central African countries, such as Benin, Burkina Faso, Senegal, and the DRC, had lower HIVST uptake. In these countries, HIVST strategies have primarily focused on key populations. In Benin, Burkina Faso, and Senegal, HIVST was largely financed by the Global Fund, and distribution prioritized to MSM and female sex workers [43]. In the DRC, HIVST was funded by PEPFAR and includes some facility-based distribution –but the emphasis remained on key populations– and is also offered through index testing and partner-delivered strategies among sero-discordant couples [44,45]. The low uptake in these settings may be further threatened by the closure of *United States Agency for International Development* (USAID) supported drop-in centers for key populations because of changes to US foreign aid and PEPFAR priorities implemented in early 2025. Innovative uses of HIVST may be increasingly employed in settings with health worker shortages and limited budgets, as they offer cost savings and efficiencies; expanded access through pharmacies and client-led testing may also become more feasible and attractive under such constraints. Our pooled meta-analysis suggested that people living with untreated HIV were less likely to use HIVST compared to individuals living without HIV, with substantial between-country heterogeneity and the associated confidence interval including the null. A potential explanation lies in our inability to exclude individuals who were already aware of their HIV status but untreated. While surveys included questions about status awareness among those who are untreated, previous research has documented high levels of non-disclosure of previous diagnosis [46]. PLHIV who are aware of their status but not on treatment may also be less inclined to self-test or disclose self-testing due to stigma or perceived lack of need.

Our study results should be interpreted considering the following limitations. First, some of the model parameters are best informed in settings with a good temporal overlap between survey and program data. However, few countries met this criterion, which led to imprecise estimates of the HIVST re-testing rate ratio and, to a lesser extent, the proportion of distributed HIVST kits used. Second, survey data on HIVST use is self-reported and could be subject to recall bias and/or misclassification. For instance, individuals might misinterpret provider-initiated rapid testing as self-testing. Third, we excluded six countries that did not have any HIVST program data. Countries that did not collect information on HIVST distribution could have lower HIVST usage, which would lead to overestimation of uptake in our aggregated estimates compared to the whole of the African region. Finally, we did not model HIVST positivity and awareness of HIV status because most surveys did not collect HIV serostatus, and the HTS program data for most countries do not report confirmatory testing following a reactive HIVST.

Strengths of our study included assembling one of the most comprehensive datasets on HIVST uptake from population-based surveys and HIVST program data from African countries. Second, we calibrated the model to all 27 countries simultaneously in a Bayesian framework, improving estimation of important parameters in data sparse settings. Third, our model sheds light on HIVST behaviors through the synthesis and triangulation of two complementary data sources [47].

## Conclusion

Our study provides evidence that HIVST is no longer a marginal HIV testing modality. Given the increasing use of HIVST, countries should continue measuring the uptake of this critical tool to understand gaps in coverage. Moving forward, epidemiological models that intend to estimate awareness of HIV status should incorporate this modality (at least in countries with moderate levels of HIVST use) for evaluating the progress towards achieving the ‘first 95’ target [48]. However, limited information exists on how HIVST are being implemented across countries and over time, which poses challenges to understand the impacts of this modality. Moreover, after the sudden dismantlement of major survey initiatives (such as DHS) in February 2025, countries will increasingly rely on other multi-country survey initiatives (i.e., MICS) or smaller country-specific surveys that may be conducted less frequently, increasing our reliance on routinely collected program data on HIVST distribution [49]. Our modeling framework has the potential to strengthen the interpretability and usability of such program data.

Our findings can support countries in optimizing the use of HIVST by prioritizing individuals most likely to be undiagnosed, thus sustaining HTS amid shifting data landscapes and funding priorities. Our modeling framework offers a practical tool for national HIV programs to monitor HIVST uptake, and guide policy decisions and interventions aimed at increasing HIV testing coverage and improving linkage to care to, ultimately, reduce HIV incidence.

## Acknowledgements

The authors acknowledge and thank all the collaborators, the survey participants and the field workers involved in the project. OE and JWI-E acknowledge funding from the MRC Centre for Global Infectious Disease Analysis (reference MR/X020258/1), funded by the UK Medical Research Council (MRC). This UK-funded award is carried out in the frame of the Global Health EDCTP3 Joint Undertaking. JWE-E acknowledges funding from the Gates Foundation [INV-005576]. The conclusions and opinions expressed in this work are those of the author(s) alone and shall not be attributed to the Foundation.

## Data Availability Statement

The individual-level data used in this study are publicly available from multiple nationally representative, population-based surveys across 27 African countries. The individual-level data from the population-based surveys included in our analyses in our analyses are publicly available and can be obtained from the following sources upon registration/request:

- Demographic and Health Surveys (DHS): https://dhsprogram.com/Data/
- Population-based HIV Impact Assessment (PHIA): https://phia-data.icap.columbia.edu/datasets
- Multiple Indicator Cluster Surveys (MICS): https://mics.unicef.org/surveys
- Kenya AIDS Indicator Survey (KAIS): https://statistics.knbs.or.ke/nada
- Botswana AIDS Indicator Survey (BAIS): https://microdata.statsbots.org.bw

Detailed information on the surveys included is provided in *Supplementary Materials, Section S1, Table S1.1*. A cleaned, survey-adjusted aggregate dataset (by age and sex), along with the analysis and modeling code is available on GitHub: https://github.com/pop-health-mod/hivst.

## Author contribution statement

MM-G and AA conceptualized the model and conceived the study. AA conducted the literature review, searched for surveys, harmonized survey data, and curated triangulated data. OE and AA reviewed and harmonized the program data. MM-G, AA, and CMD developed and refined the model for trend estimation. All the authors contributed to developing the methodology, quality assurance and provided inputs for the analyses. AA performed statistical analyses. AA, MM-G, and CMD drafted the manuscript. All authors critically reviewed the manuscript, contributed important intellectual content, and approved the final version for submission. The findings and conclusions in this report are those of the authors and do not necessarily represent the official position of the funding agencies.

## Prior posting and presentation

This work is the sole product of the authors and has never been submitted for publication. A preprint will be posted shortly.

